# Association of cerebral small vessel disease burden with brain structure and cognitive and vascular risk trajectories in mid-to-late life

**DOI:** 10.1101/2021.04.13.21255391

**Authors:** Michelle G Jansen, Ludovica Griffanti, Clare E Mackay, Melis Anatürk, Luca Melazzini, Ann-Marie G de Lange, Nicola Filippini, Enikő Zsoldos, Kim Wiegertjes, Frank-Erik de Leeuw, Archana Singh-Manoux, Mika Kivimäki, Klaus P Ebmeier, Sana Suri

## Abstract

We characterize the associations of total cerebral small vessel disease (SVD) burden with brain structure, trajectories of vascular risk factors, and cognitive functions in mid-to-late life. Participants were 623 community-dwelling adults from the Whitehall II Imaging Sub-study with multi-modal MRI (mean age 69.96 SD=5.18, 79% men). We used linear mixed-effects models to investigate associations of SVD burden with up to 25-year retrospective trajectories of vascular risk and cognitive performance. General linear modelling was used to investigate concurrent associations with grey matter (GM) density and white matter (WM) microstructure, and whether these associations were modified by cognitive status (Montreal Cognitive Assessment, MoCA). Severe SVD burden in older age was associated with higher mean arterial pressure throughout midlife (β=3.36, 95% CI [0.42-6.30]), and faster 25-year cognitive decline in letter fluency (β=-0.07, 95% CI [-0.13–-0.01]), and verbal reasoning (β=-0.05, 95% CI [-0.11–-0.001]). Moreover, SVD burden was related to lower GM volumes in 9.7% of total GM, and widespread WM microstructural decline (FWE-corrected *p*<0.05). The latter association was most pronounced in individuals with cognitive impairments on MoCA (*F*_*3,608*_=2.14, *p*=0.007). These findings highlight the importance of managing midlife vascular health to preserve brain structure and cognitive function in old age.

## Introduction

Cerebral small vessel disease (SVD) refers to a series of pathological processes, damaging the small perforating arterioles of the brain.^1^ Conventional MRI markers of SVD include, amongst others, white matter hyperintensities (WMH), enlarged perivascular spaces (EPVS), cerebral microbleeds (CMB), and lacunes.^1, 2^ The total SVD burden score combines the visual ratings of these markers, providing a more comprehensive measure of SVD than each individual component alone.^3, 4^

Accumulating evidence suggests that controlling vascular risk factors (e.g., blood pressure, obesity) earlier in the lifespan may preserve structural brain health in older ages and hence contribute to preventing dementia.^5-9^ However, previous research investigating longitudinal correlates of the SVD burden score have been limited to stroke patients or solely examined sociodemographic factors (e.g., intelligence at age 11).^10, 11^ SVD burden varies considerably between healthy older adults^10^ and contributes to several other brain structural impairments.^1^ However, the relationship between total SVD scores and brain morphology is less clear.^12^ Studies investigating the longitudinal vascular and cognitive correlates of SVD, together with structural brain associations, may provide further insights into the stage in life when interventions designed to promote healthy aging could ideally be administered.

We investigated the associations of total SVD burden with up to (1) 25-year retrospective trajectories of vascular risk factors (mean arterial pressure [MAP]; body mass index [BMI]; Framingham Stroke Risk Score [FSRS]) and cognitive decline on several domains and (2) concurrent grey matter (GM) density and white matter (WM) microstructure in 623 elderly individuals from the Whitehall II Imaging cohort. We also examined whether the associations of SVD burden with brain structure were moderated by cognitive status in older age.

## Methods

### Study design and participants

Participants were selected from the Whitehall II Imaging cohort, a sub-study of the prospective Whitehall II cohort, established by University College London in 1985. Whitehall II participants have received detailed clinical follow-ups for up to 30 years in 1991-1994 (Wave 3), 1997-1999 (Wave 5), 2002-2004 (Wave 7), 2007-2009 (Wave 9), 2012-2013 (Wave 11), and 2015-2016 (Wave 12). For the Imaging Sub-study, 800 participants (60-85 years old) were randomly selected from the Whitehall II Wave 11 cohort and underwent a detailed neuropsychological assessment and multi-modal 3T brain MRI scans at the University of Oxford between 2012-2016 (MRI Wave).^13^ A total of 623 participants were included after removing participants with missing data for variables of interest in three or more waves (N=36), the occurrence of gross MRI abnormalities (e.g., large strokes, cysts, tumours, hydrocephalus which failed the MRI pre-processing pipelines; N=28), and missing or insufficient quality of MRI images for analysis (e.g., motion artefacts; N=88). A detailed description and flowchart of participant inclusion is presented in **Supplementary Methods and Figure S1**. The Whitehall II cohort profile and the Imaging Sub-study protocol have been described previously.^13, 14^

### Standard protocol approvals, registrations, and patient consents

The University of Oxford Central University Research Ethics Committee (reference: MS IDREC-C1-2011-71) and the University College London Medical School Committee on the Ethics of Human Research (reference: 85/0938) provided ethical approval for the study. Informed and written consent was given by each participant enrolled in the study at each wave.

### Demographics

Demographics included age, sex, Caucasian ethnicity, years of full-time education (self-reported at MRI Wave), and highest employment grade at Wave 3 in 1991-1994 to indicate socio-economic status (high=managers/administrators, intermediate=professionals/executives, low=clerical/support staff).

### Neuroimaging

MRI scans were acquired on a 3T Siemens Magnetom Verio scanner (Erlangen, Germany) between April 2012–December 2014 (N=427). After a scanner upgrade, the remaining 196 scans were acquired on a 3T Siemens Magnetom Prisma scanner (Erlangen, Germany) between June 2015–April 2016. We examined the following sequences: high-resolution T1-weighted images; fluid-attenuated recovery imaging (FLAIR); T2*-weighted images; and diffusion weighted imaging. Sequences were closely matched between the two scanners and acquisition parameters are described in **Table S1**, and elsewhere.^6, 13^

### Total SVD burden

Visual ratings were performed by experienced raters in accordance with the Standards for Reporting Vascular Changes on Neuroimaging (STRIVE) criteria.^2^ Periventricular and deep WMH were rated on FLAIR images, lacunes on T1 and FLAIR images, EPVS on T1 images, and CMB on T2* images. Details on visual ratings and intra-rater reliability are provided in **Supplementary Methods**. These ratings were used to calculate total SVD scores to express SVD severity on an ordinal scale from 0-4; 1 point each was provided for a Fazekas score of deep WMH≥2 and/or periventricular WMH>2, CMB count≥1, lacunes≥1, and EPVS≥11 in the basal ganglia of one hemisphere.^3, 4^ As only 2% of the sample (N=13) had a SVD burden score of 4, we merged groups with scores of 3 and 4.

### Vascular risk factors

Vascular risk factors were assessed six times at 5-year intervals from 1991-1994 (Wave 3) to 2015-2016 (Wave 12), using both questionnaires and clinical examinations (for details, see **Supplementary Methods)**. Based on the literature linking vascular risk factors and cognitive decline,^6-8^ we selected three measures of vascular risk for the longitudinal trajectory analysis: MAP [(systolic BP + 2 x diastolic BP)/3], BMI (kg/m^2^) and the FSRS. The FSRS predicts the likelihood of a stroke within 10 years, calculated with an algorithm which combines age, sex, systolic blood pressure, self-reported use of antihypertensives, diabetes, current smoking, current or previous atrial fibrillation, left ventricular hypertrophy, and cardiovascular disease.^15^

At the MRI Wave, weekly alcohol consumption was self-reported, and metabolic equivalents of task (METs) per week for moderate-to-vigorous activity was calculated using the self-administered Community Healthy Activities Model Program for Seniors (CHAMPS) questionnaire.^16^

### Cognitive function

Cognitive function was evaluated five times at 5-year intervals from 1997-1999 (Wave 5) to 2015-2016 (Wave 12), covering letter fluency (number of ‘S’ words listed in one minute), semantic fluency (category ‘animals’), short-term memory (recall of a list of 20 words in two minutes), and verbal and numerical reasoning as assessed with the Alice Heim 4-I test.^17^

We also used a single measure of Montreal Cognitive Assessment (MoCA), evaluated only at the MRI Wave, to assess cognitive status outcome. We classified cognitive impairments as performance below the traditional established screening cut-offs (MoCA scores<26).^18^ Detailed information on the above measures is provided in **Supplementary Methods**.

### MRI Outcomes

MRI scans were analysed using FMRIB Software Library v6.0 (FSL; https://fsl.fmrib.ox.ac.uk/). Detailed pre-processing pipelines have been described in the protocol paper^13^ and in **Supplementary Methods**. Briefly, pre-processed T1 images were analysed using voxel-based morphometry (FSL-VBM) to produce GM density maps for each participant. Pre-processing of DTI images produced four maps for each participant: fractional anisotropy (FA), mean diffusivity (MD), radial diffusivity (RD), and axial diffusivity (AD), which are widely used to estimate axonal and myelin integrity.^19^ We used Tract-Based Spatial Statistics (TBSS) to create a skeletonized (thinned) FA, MD, RD and AD maps. All maps were concatenated in 4D files for subsequent voxel-wise statistics. Measures of mean global FA, MD, RD and AD were extracted from the respective mean skeletonized maps. WMH were segmented on FLAIR images using the supervised FMRIB’s Brain Intensity AbNormality Classification Algorithm (BIANCA) tool.^20^ Binarized WMH maps were generated by selecting voxels that exceeded a probability of 0.9 of being WMH on the BIANCA output, and used to quantify WMH volumes as % of total brain volume.

### Statistical analysis

All statistical analyses were performed using R version 3.6.1. Effects were considered significant when *p*< 0.05 (two-tailed). Participant characteristics and physiological measurements were compared between SVD groups (0-3) using one-way analysis of variance (ANOVA), Kruskall-Wallis test, and Chi-square test where appropriate. Effect sizes were calculated where possible and expressed with Cohen’s f^2^ (f^2^=0.02 small; medium f^2^=0.15; f^2^=0.35 large).

Linear mixed effect models (LME) with random slopes and intercepts were used to investigate whether individuals in the four SVD burden groups differed in baseline measures and longitudinal trajectories of vascular risk factors (MAP, BMI, FSRS) and cognition (letter and categorical fluency, memory, verbal and numerical reasoning). A complete description of the LME models is provided in **Supplementary Methods**. FSRS was log-transformed due to the skewed distribution of the standardized residuals. A binary covariate was added to control for the effects of MRI scanner model. Further, we corrected for the effects of age, sex, and education, by scaling these variables and incorporating their main effects and interactions with the time terms. A linear model best described MAP and BMI trajectories; however, including an orthogonalized polynomial quadratic time term improved model fit for FSRS and cognitive trajectories (all *p*<.05).^21, 22^ This quadratic time term expresses non-linear (exponential) changes in the LME model without inducing collinearity.^23^ We performed Bonferroni corrections for multiple comparisons across the 5 cognitive measures. Accordingly, results surpassing a strict significance threshold (*p*<0.01) are discussed in detail whereas those 0.01< *p*<0.05 are interpreted with caution.

Voxel-wise associations of SVD burden with GM density and DTI-derived metrics (FA, MD, RD, AD) were investigated using general linear modelling (GLM) and permutation-based non-parametric testing (5000 permutations) using the FSL Randomize Tool. A separate voxel-wise GLM was performed for each DTI metric, and included the covariates of no interest: age, sex, education, MRI scanner model, antihypertensive use, BMI, and MAP measured at the time of MRI. Analyses were corrected for multiple voxel-wise comparisons using family-wise error (FWE) corrections and reporting threshold-free cluster enhancement (TFCE) statistics. We applied a Bonferroni correction for multiple comparisons across the 4 DTI metrics, accepting a strict significance threshold of *p*<0.0125. To examine whether the effects of SVD burden on DTI metrics were driven by the presence of WMH, we performed a sensitivity analysis which included the WMH spatial maps as voxel-wise confounds in the model.

We also examined whether the associations of SVD burden with global measures of GM and WM microstructure were moderated by cognitive status (MoCA<26 vs. MoCA≥26) at the time of MRI. Multivariate analysis of covariance (MANCOVA) was performed with cognitive status and SVD burden as independent variables, and global GM, mean global FA, MD, RD, and AD as dependent variables, and the same covariates as above.

### Data availability

The study follows Medical Research Council data sharing policies (https://mrc.ukri.org/research/policies-and-guidance-for-researchers/data-sharing), and data from the Whitehall II Study and Whitehall II Imaging Sub-study are accessible via application to the Dementias Platform UK (https://portal.dementiasplatform.uk).

## Results

At the time of MRI, the mean age of the 623 participants was 69.96 years (SD=5.18), 494 (79%) were men and 594 (95%) Caucasian, reflecting the demographics of the parent Whitehall II study. Sample demographics, physiological measurements, and the distribution of the total SVD score at the time of MRI (2012-2016) are presented in **Table 1**.

**Table 1.**
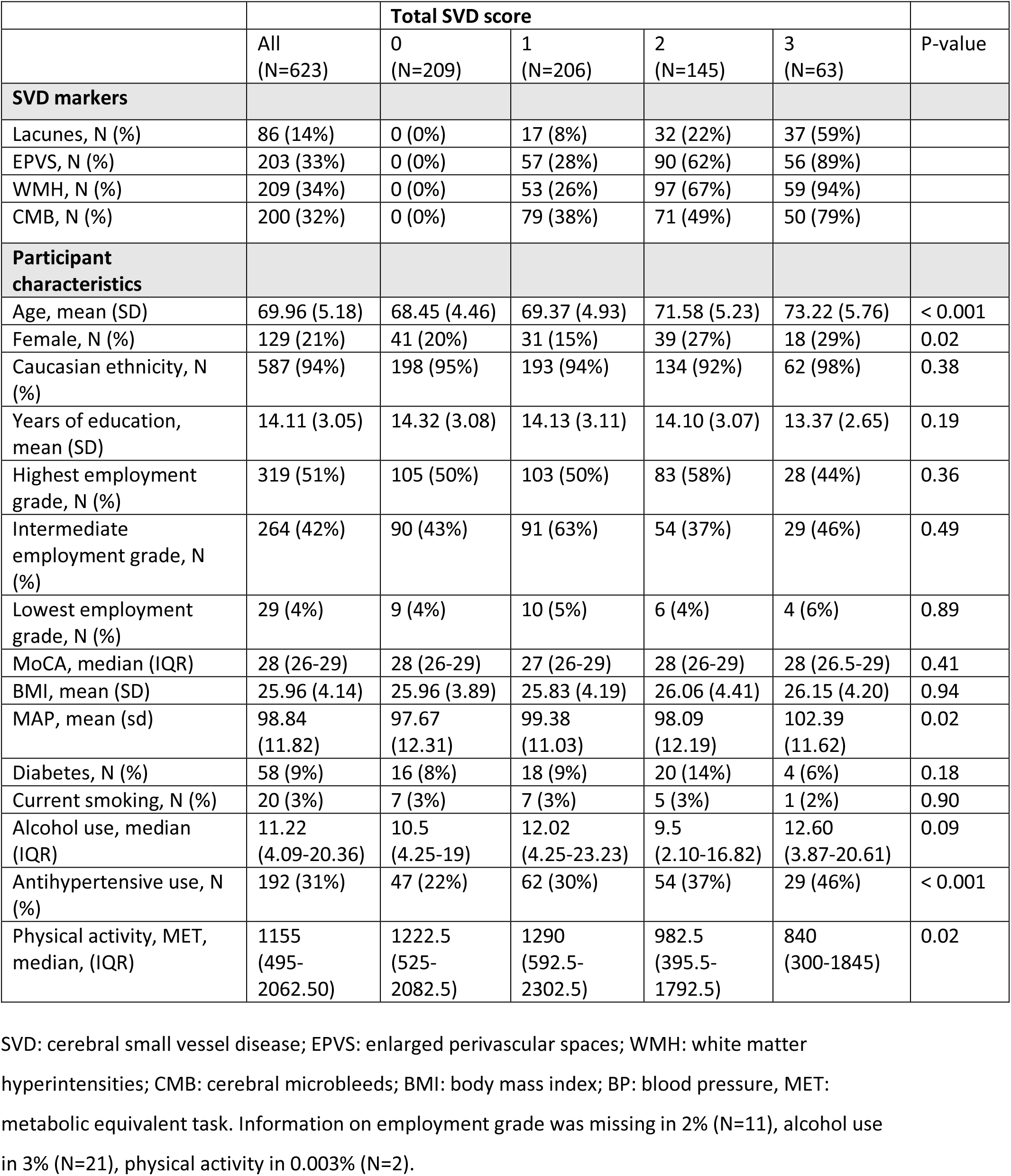
Participant characteristics and associations at the MRI wave (2012-2016).

|  |  | Total SVD score |  |  |  |  |
| --- | --- | --- | --- | --- | --- | --- |
|  | All<br>(N=623) | 0<br>(N=209) | 1<br>(N=206) | 2<br>(N=145) | 3<br>(N=63) | P-value |
| **SVD markers** |  |  |  |  |  |  |
| Lacunes, N (%) | 86 (14%) | 0 (0%) | 17 (8%) | 32 (22%) | 37 (59%) |  |
| EPVS, N (%) | 203 (33%) | 0 (0%) | 57 (28%) | 90 (62%) | 56 (89%) |  |
| WMH, N (%) | 209 (34%) | 0 (0%) | 53 (26%) | 97 (67%) | 59 (94%) |  |
| CMB, N (%) | 200 (32%) | 0 (0%) | 79 (38%) | 71 (49%) | 50 (79%) |  |
| **Participant characteristics** |  |  |  |  |  |  |
| Age, mean (SD) | 69.96 (5.18) | 68.45 (4.46) | 69.37 (4.93) | 71.58 (5.23) | 73.22 (5.76) | < 0.001 |
| Female, N (%) | 129 (21%) | 41 (20%) | 31 (15%) | 39 (27%) | 18 (29%) | 0.02 |
| Caucasian ethnicity, N (%) | 587 (94%) | 198 (95%) | 193 (94%) | 134 (92%) | 62 (98%) | 0.38 |
| Years of education, mean (SD) | 14.11 (3.05) | 14.32 (3.08) | 14.13 (3.11) | 14.10 (3.07) | 13.37 (2.65) | 0.19 |
| Highest employment grade, N (%) | 319 (51%) | 105 (50%) | 103 (50%) | 83 (58%) | 28 (44%) | 0.36 |
| Intermediate employment grade, N (%) | 264 (42%) | 90 (43%) | 91 (63%) | 54 (37%) | 29 (46%) | 0.49 |
| Lowest employment grade, N (%) | 29 (4%) | 9 (4%) | 10 (5%) | 6 (4%) | 4 (6%) | 0.89 |
| MoCA, median (IQR) | 28 (26-29) | 28 (26-29) | 27 (26-29) | 28 (26-29) | 28 (26.5-29) | 0.41 |
| BMI, mean (SD) | 25.96 (4.14) | 25.96 (3.89) | 25.83 (4.19) | 26.06 (4.41) | 26.15 (4.20) | 0.94 |
| MAP, mean (sd) | 98.84 (11.82) | 97.67 (12.31) | 99.38 (11.03) | 98.09 (12.19) | 102.39 (11.62) | 0.02 |
| Diabetes, N (%) | 58 (9%) | 16 (8%) | 18 (9%) | 20 (14%) | 4 (6%) | 0.18 |
| Current smoking, N (%) | 20 (3%) | 7 (3%) | 7 (3%) | 5 (3%) | 1 (2%) | 0.90 |
| Alcohol use, median (IQR) | 11.22 (4.09-20.36) | 10.5 (4.25-19) | 12.02 (4.25-23.23) | 9.5 (2.10-16.82) | 12.60 (3.87-20.61) | 0.09 |
| Antihypertensive use, N (%) | 192 (31%) | 47 (22%) | 62 (30%) | 54 (37%) | 29 (46%) | < 0.001 |
| Physical activity, MET, median, (IQR) | 1155 (495-2062.50) | 1222.5 (525-2082.5) | 1290 (592.5-2302.5) | 982.5 (395.5-1792.5) | 840 (300-1845) | 0.02 |

SVD: cerebral small vessel disease; EPVS: enlarged perivascular spaces; WMH: white matter hyperintensities; CMB: cerebral microbleeds; BMI: body mass index; BP: blood pressure, MET: metabolic equivalent task. Information on employment grade was missing in 2% (N=11), alcohol use in 3% (N=21), physical activity in 0.003% (N=2).

Individuals with higher SVD burden scores were older, more often female, demonstrated higher MAP, were more likely to be on antihypertensive treatment, and engaged in less physical activity. Participants’ demographics and vascular risk factors at the five follow-up waves are shown in **Table S2**. The average follow-up time from Wave 3 to Wave 12 was 23.5 years (SD=0.57, interquartile range [IQR] 23.17-23.91). The time from Wave 3 to the MRI was on average 22.15 years (SD=1.40, IQR 21.02-23.39), and from Wave 5 to MRI was 16.38 years (SD=1.30, IQR 15.27-17.56). The demographic characteristics of the 623 included participants did not differ from the starting sample of N=775 with scans from the Whitehall II Imaging Sub-Study **(Table S3)**. Thirty-four percent of the sample demonstrated total SVD scores of 0, followed by scores of 1 (33%), 2 (23%) and 3 (10%). Lacunes were the least frequently observed SVD feature (14%), whereas EPVS, WMH and CMB were all equally prevalent (32-34%; see **Figure S2** for a Venn-diagram).

Relative to the reference SVD burden group (SVD=0), the highest SVD burden group had higher baseline MAP (measured approximately 22.15 years prior to the MRI Wave at mean age=47.81; SD=5.23; *p*=0.03, Cohen’s f^2^=0.01) after correcting for age, sex, education, and MRI scanner model. However, the SVD groups did not differ in baseline BMI and FSRS, or rates of change of MAP, BMI and FSRS over the 30-year follow up period (**Table 2; Figure 1**).

**Table 2.** Longitudinal associations of vascular factors with SVD burden.

|  | MAP |  | BMI |  | Log-FSRS |  |
| --- | --- | --- | --- | --- | --- | --- |
|  | Beta | 95% CI | Beta | 95% CI | Beta | 95% CI |
| **Main effect of SVD burden group** |  |  |  |  |  |  |
| SVD 1 vs. 0 | 1.75 | -0.18 to 3.67 | 0.04 | -0.63 to 0.70 | 0.06 | -0.002 to 0.11 |
| SVD 2 vs. 0 | 1.39 | -0.79 to 3.57 | 0.06 | -0.70 to 0.82 | 0.05 | -0.02 to 0.11 |
| SVD 3 vs. 0 | 3.36 | 0.42 to 6.30<sup>a</sup> | 0.13 | -0.89 to 1.15 | 0.05 | -0.03 to 0.14 |
| **Interaction SVD group and time** |  |  |  |  |  |  |
| Time*SVD 1 vs. 0 | -0.02 | -0.13 to 0.08 | 0.01 | -0.03 to 0.01 | 0.001 | -0.002 to 0.005 |
| Time*SVD 2 vs. 0 | -0.05 | -0.17 to 0.06 | 0.002 | -0.02 to 0.02 | 0.001 | -0.002 to 0.01 |
| Time*SVD 3 vs. 0 | -0.06 | -0.21 to 0.10 | -0.004 | -0.03 to 0.03 | 0.002 | -0.003 to 0.01 |
| **Interaction SVD group and time<sup>2</sup>** |  |  |  |  |  |  |
| Time<sup>2</sup>*SVD 1 vs. 0 |  |  |  |  | -0.02 | -0.04 to 0.003 |
| Time<sup>2</sup>*SVD 2 vs. 0 |  |  |  |  | -0.01 | -0.04 to 0.01 |
| Time<sup>2</sup>*SVD 3 vs. 0 |  |  |  |  | 0.001 | -0.03 to 0.04 |

Results of linear mixed effect models with random effects for the intercept and slope (time for MAP, BMI FSRS and time<sup>2</sup> for FSRS). Vascular risk factors were assessed between Wave 3 (1991-1994) and Wave 12 (2015-2016). All models were adjusted for age at baseline, sex, education, and scanner.
MAP: mean arterial pressure; BP: blood pressure; FSRS: Framingham Stroke Risk Score; CI: confidence interval; SVD: cerebral small vessel disease. <sup>a</sup> $p < 0.05$ .

**Figure 1.**
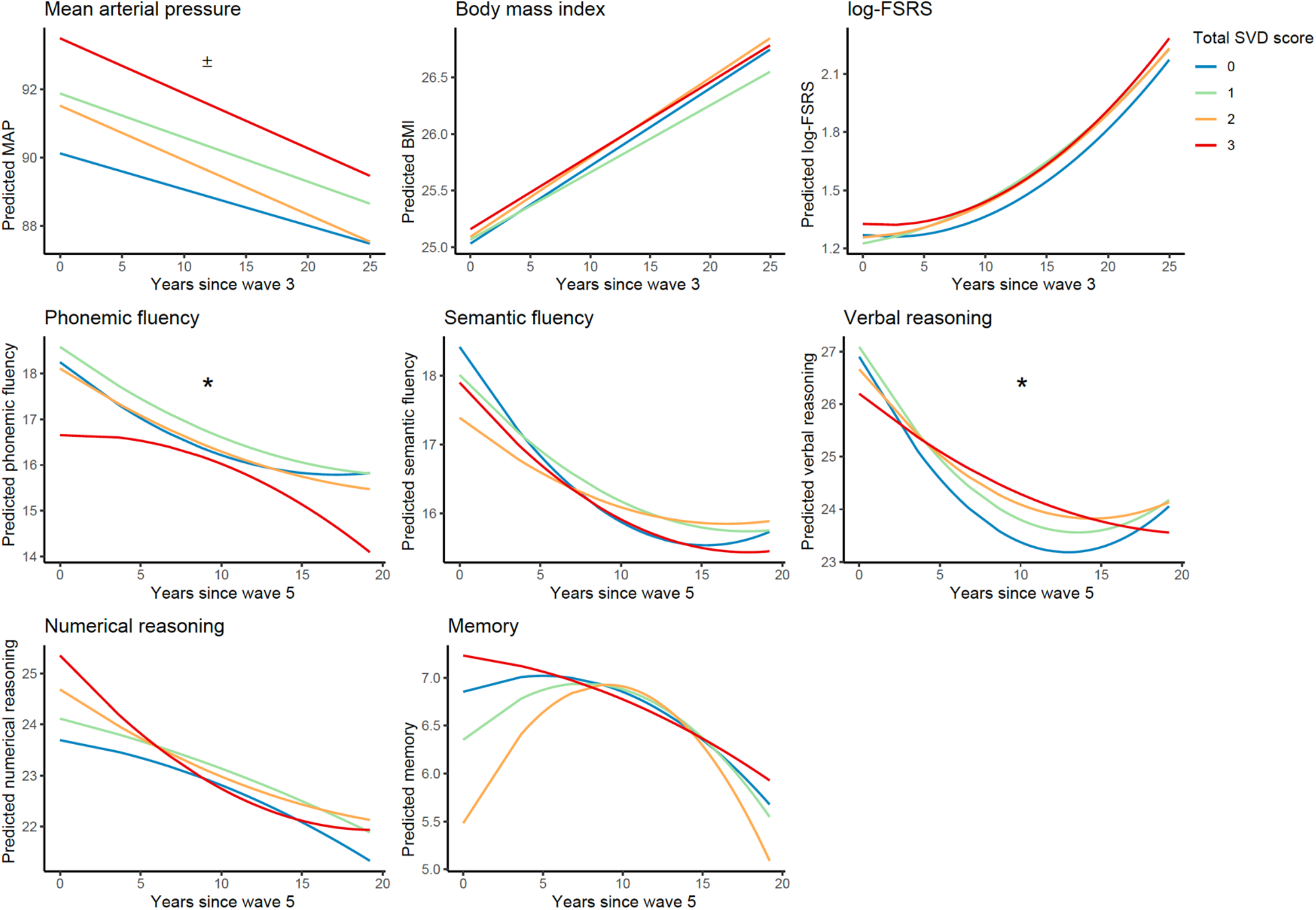
Predicted longitudinal trajectories for vascular risk and cognitive performance. Each figure depicts the predicted trajectories of the dependent variable of interest based on the linear mixed effect models. Measures for mean arterial pressure (MAP) and log-Framingham stroke risk score (log-FSRS) were obtained between 1995 and 2016. Measures on cognitive performance were obtained between 1997 and 2016. ± indicates significant main effects of SVD burden, where group 3 had higher baseline values compared to group 0 (*p*<0.05). * indicates significant interactions where SVD burden group 3 showed steeper rates of cognitive decline compared to group 0 (*p*<0.05). MAP: mean arterial pressure. BMI: body mass index; FSRS: Framingham Stroke Risk Score; SVD: cerebral small vessel disease.

After adjustments for age, sex, education and MRI scanner model, relative to the reference SVD group (SVD=0), individuals with higher SVD burden had steeper linear declines in letter fluency (*p* =0.02, Cohen’s f^2^=0.002) as well as linear (*p*=0.047, Cohen’s f^2^=0.002) and exponential (*p*=0.04, Cohen’s f^2^=0.002) declines in verbal reasoning. However, these associations did not survive Bonferroni corrections. The SVD burden groups did not differ on baseline cognitive performance or rate of cognitive decline for letter fluency, and verbal and numerical reasoning (summary statistics in **Table 3; Figure 1**).

**Table 3.**
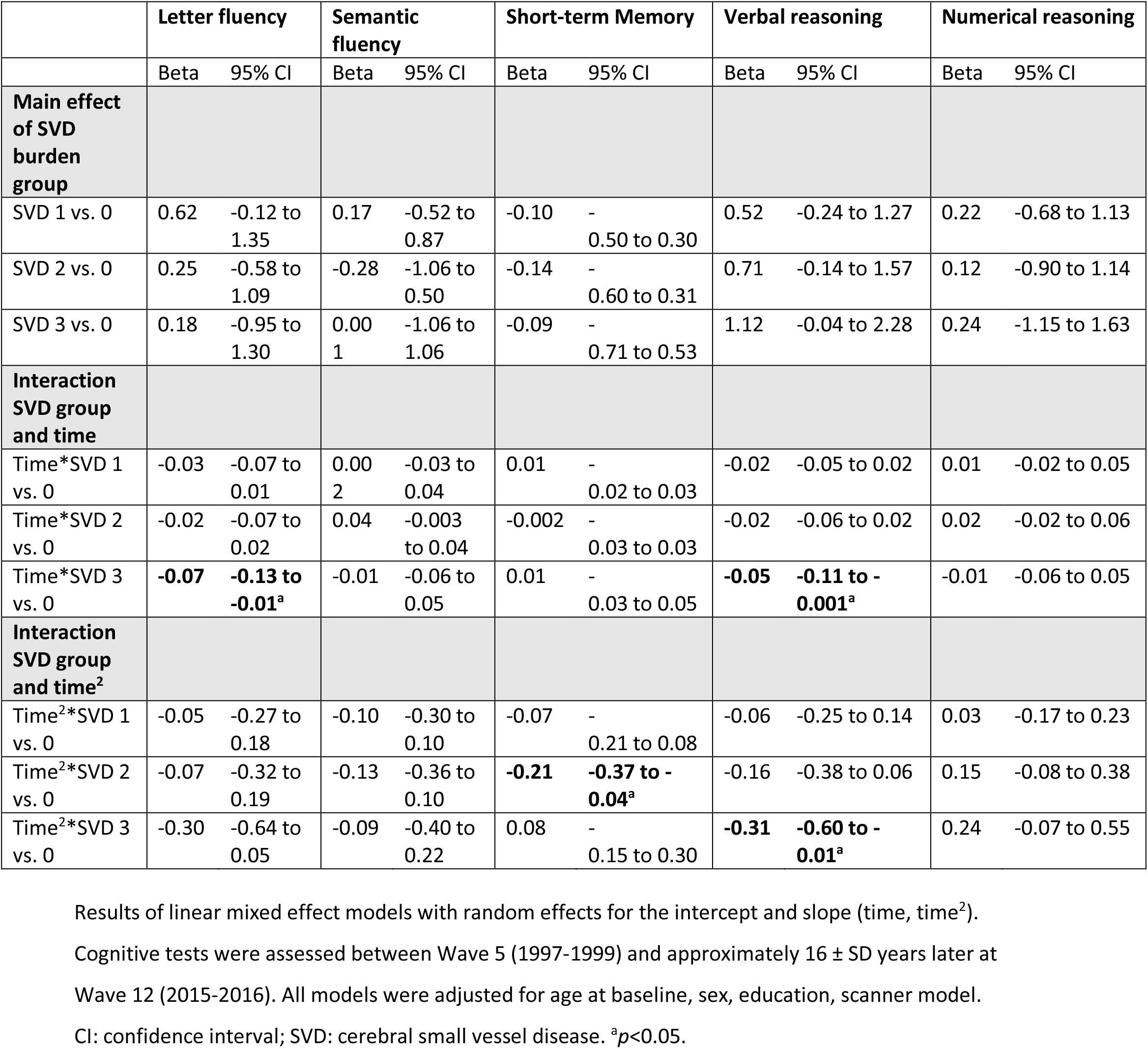
Longitudinal associations of cognitive performance with SVD burden.

VBM analysis revealed that higher SVD burden was associated with lower GM density in several areas, including the frontal pole, superior and inferior frontal gyrus, pre- and postcentral gyrus, precuneus, frontal orbital cortex, subcallosal cortex, left Heschl’s gyrus, parahippocampal gyrus, and hippocampus (**Figure 2**, FWE-corrected *p*<0.05). Together, these regions covered 9.7% of the total brain GM volume.

**Figure 2.**
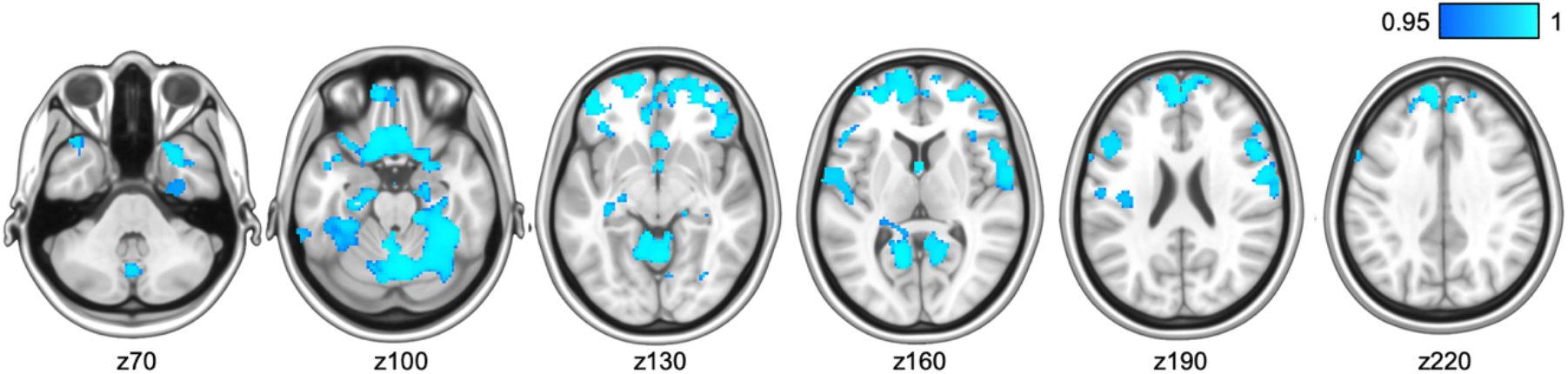
Voxel-wise associations between higher total cerebral small vessel disease (SVD) scores and lower grey matter density were primarily found cortical and hippocampal areas, shown in blue. TFCE-corrected statistical maps are overlaid on the MNI-152 template and thresholded at a 1-p value of 0.95, representing *p*<0.05. Images are also FWE-corrected for multiple voxel-wise comparisons. Analysis was adjusted for age, sex, education, MRI scanner model, antihypertensive use, body mass index, and mean arterial pressure measured at the time of the MRI scan.

Voxel-wise TBSS analyses revealed that higher SVD MRI burden was associated with lower FA, and higher MD, RD, and AD (Bonferroni-corrected *p*<0.0125) throughout several projection and commissural WM tracts such as the corpus callosum, cingulum, corona radiata, and longitudinal fasciculus (**Figure 3**). Associations were distributed across 17.0%, 16.7%, 15.4% and 23.8% of total WM tract skeletons for FA, MD, RD and AD, respectively. These results remained after a voxel-wise correction for WMH masks.

**Figure 3.**
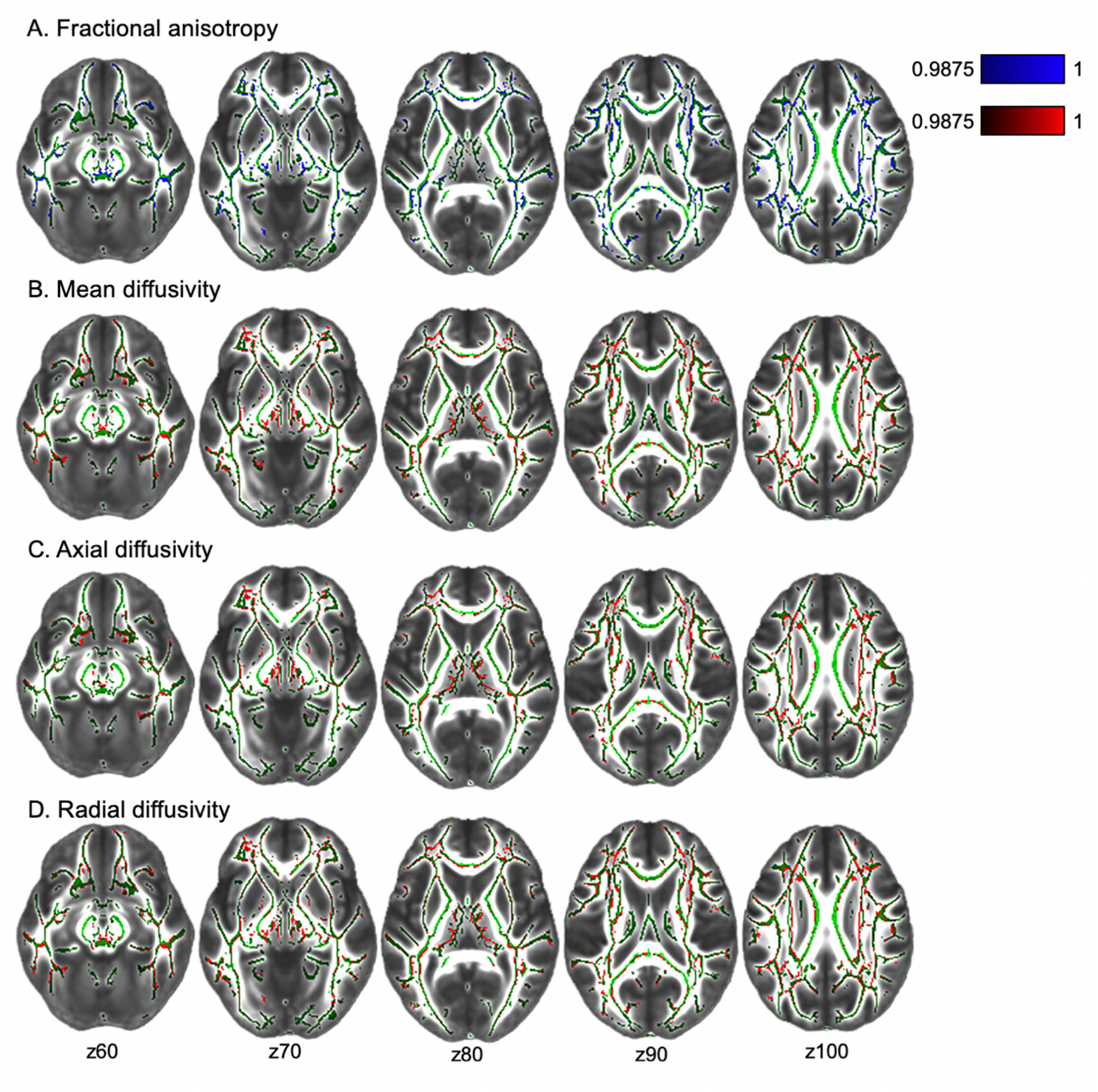
Voxel-wise associations between cerebral small vessel disease (SVD) burden and diffusion tensor imaging measures: fractional anisotropy (FA; A) mean diffusivity (MD; B); axial diffusivity (AD; C), and radial diffusivity (RD; D). Statistical maps were overlaid on the FMRIB58_FA standard image. Green tracts depict the standardized mean FA skeleton. Higher SVD burden is associated with lower FA (in blue), and higher MD, AD, RD (in red). All images are TFCE-corrected statistics thresholded at 1-p values of 0.9875, i.e. representing Bonferroni-corrected *p*<0.0125 to adjust for multiple comparisons across 4 DTI metrics. Images are also FWE-corrected for multiple voxel-wise comparisons. Analyses were adjusted for age, sex, education, MRI scanner model, antihypertensive use, body mass index, and mean arterial pressure measured at the time of the MRI scan.

We observed an interaction between SVD burden and cognitive status (*F*_*3,608*_=2.14, *p*=0.007, Cohen’s f^2^=0.13). Post-hoc univariate comparisons revealed that this interaction effect was driven by MD (*F*_*3,608*_=3.27, *p*=0.02, Cohen’s f^2^=0.13), AD (*F*_*3,608*_=3.72, *p*=0.01, Cohen’s f^2^=0.14), and RD (*F*_*3,608*_ =2.72, *p*=0.04, Cohen’s f^2^=0.12), but not FA (*F*_*3,608*_ =1.03, *p*=0.38) or GM (*F*_*3,608*_ =0.97, *p*=0.41). For each of these interactions, the association of SVD burden with poor WM microstructure was more pronounced in individuals with cognitive impairments (reflected by MoCA<26, **Figure 4**).

**Figure 4.**
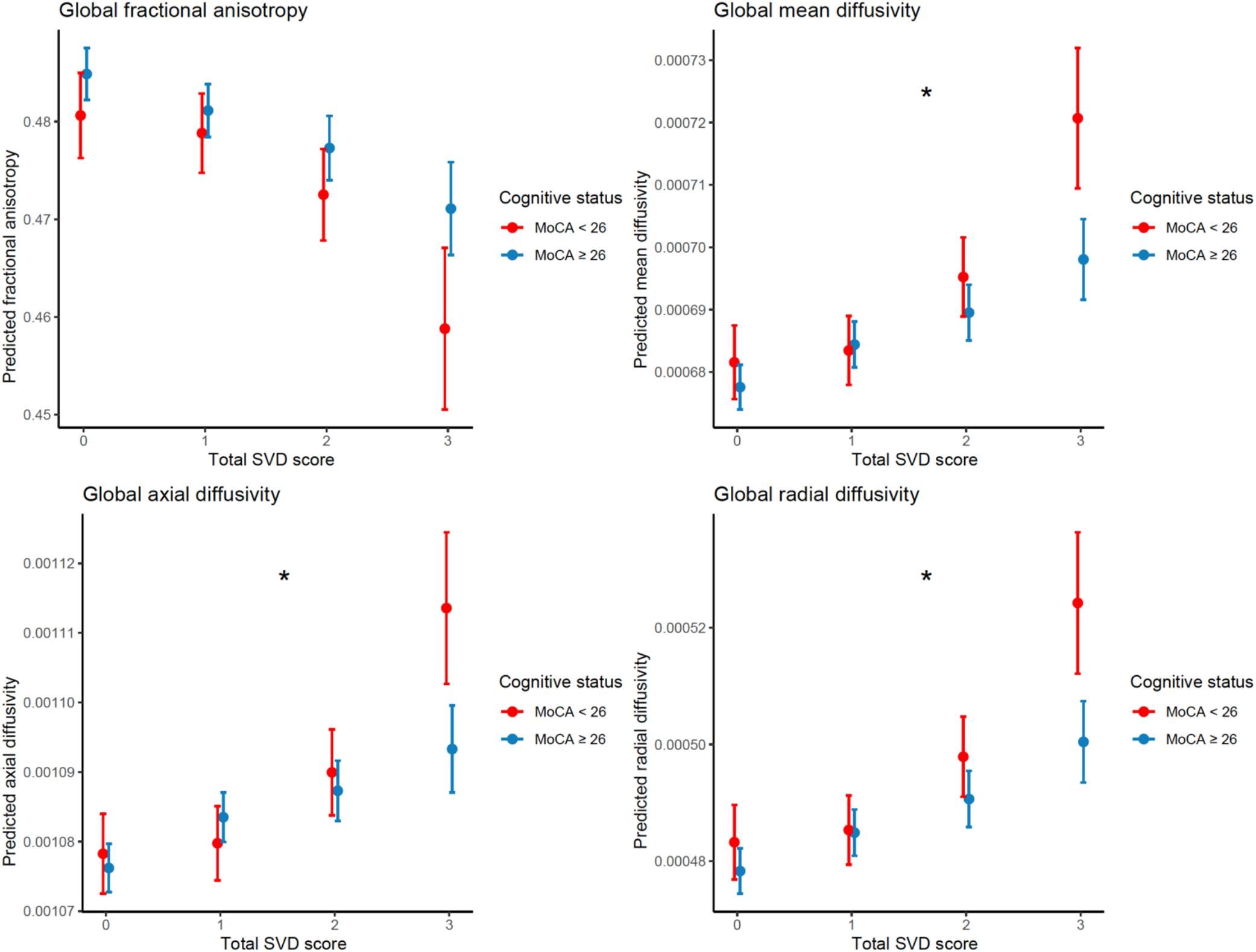
Plots depict significant interactions between cognitive status (Montreal Cognitive Assessment [MoCA]<26 vs. ≥ 26) and total SVD scores (0-3) on mean diffusivity, axial diffusivity, and radial diffusivity (all *p<*.05; depicted with *), but not for fractional anisotropy. The association between SVD burden and DTI parameters is more pronounced in individuals with cognitive impairments, reflected by MoCA<26, and especially in the most severe SVD burden group. MoCA: Montreal Cognitive Assessment; SVD: cerebral small vessel disease.

## Discussion

This study characterized late-life SVD burden across a large sample of community-dwelling older adults and has demonstrated several associations with 25-year retrospective trajectories of vascular risk factors, cognitive performance, and brain microstructure. We showed that individuals with severe SVD burden at the MRI Wave presented with elevated mean arterial pressure 25 years prior to their MRI (Wave 3). We also found that individuals with higher SVD had relatively faster rates of decline for letter fluency and verbal reasoning over this timespan. In addition, at the time of MRI, we revealed concurrent associations of SVD burden with widespread reductions in GM density as well as hallmark indicators of WM structural integrity, reflected by reduced FA, and increased diffusivity. Importantly, the negative associations between SVD and diffusivity were stronger in individuals with cognitive impairments (MoCA<26). Overall, our findings could have important implications for optimizing strategies for maintaining brain and cognitive health during the lifespan.

While previous studies have revealed associations between higher midlife blood pressure and late-life SVD in normal aging, these have only examined either WMH, CMB, or brain infarcts individually.^7, 9^ Here, we show that higher MAP in midlife (mean age 48, Wave 3) is associated with severe total SVD burden approximately 20 years later (mean age 70, MRI Wave), measured as a combination of four key MRI markers of SVD pathology.^4^ Our results thus indicate that elevated midlife MAP can have more diffuse effects on cerebrovascular health that go beyond the single associations with separate SVD features. Our findings are in line with previous studies showing that hypertension exacerbates SVD-related pathologies in aging individuals.^1, 24^ We found no evidence for the involvement of other midlife vascular risk factors (BMI or stroke risk scores) in late-life SVD burden, suggesting that midlife blood pressure may particularly aggravate SVD-related injuries and therefore may serve as the most important modifiable risk factor for SVD.^25^ This study therefore adds to the accumulating evidence emphasizing the need for early prevention strategies with a focus on blood pressure management.^5-7, 9^ However, individuals with SVD are not always hypertensive, and given that our results are observational, we cannot determine causality, or rule out reverse causation.^26^ Therefore, our findings and potential clinical consequences warrant further investigation.

WMH, infarcts, CMBs have each individually been linked to cognitive decline and dementia, although the association with EPVS is less clear.^27^ Two previous studies demonstrated steeper rates of decline of global cognition and executive function, however these links were only established in hypertensive patients^28^ and symptomatic SVD.^29^ Our findings add to this by showing for the first time that, among community-dwelling elderly without diagnosis of stroke or dementia, late-life SVD burden relates to cognitive decline on letter fluency and verbal reasoning measured over the previous 20-years. As SVD features are common in elderly individuals and increase risk of all-cause dementia, including Alzheimer’s and vascular dementia^27^, these results highlight the importance of investigating how SVD links to preclinical cognition impairments. However, we made two key observations: first, that these associations were only present in participants with ≥3 SVD features, which occurred in 10% of the total sample. Similar to an earlier study^30^, there appeared to be a threshold effect, where cognitive dysfunctions became noticeable only in those with moderate-to-severe SVD burden. Second, the effects of SVD on cognition were generally small, and did not survive strict corrections for multiple-comparison corrections. While it is possible that larger studies could confirm our observed associations at an uncorrected *p*<0.05, accumulating evidence suggests that there is a large inter-individual variability in the effects of SVD burden on cognition, both with regards to the affected domains and severity.^24, 31^ Overall we observed that despite their high SVD burden, individuals displayed relatively modest increases in earlier rates of cognitive decline. This could partially be explained by the concept of reserve, which refers to the ability to alleviate the effects of neuropathology through variations in brain structure (i.e., brain reserve) or adaptability of functional processes (i.e. cognitive reserve).^32^ Brain reserve is generally considered to be a less dynamic construct, whereas cognitive reserve is thought to be more adjustable through life experiences (e.g., by increasing cognitive engagement, physical activity, leisure activities)^32^ and therefore may serve as viable target to improve clinical management of the consequences of SVD.^8, 24^

Additionally, we found specific GM and WM correlates of current SVD-related burden. More specifically, we identified GM density reductions predominantly in the medial-frontal, orbito-frontal, and medial-temporal regions. This pattern of cortical atrophy has been recognized in earlier studies examining WMH or lacunes alone.^33, 34^ These associations were most pronounced in the medial temporal and hippocampal regions, which are traditionally linked to AD pathology.^35^ This is especially important given the growing recognition of the prevalence of SVD and cerebrovascular dysfunction in AD.^36^

Grey matter atrophy may result from secondary neurodegenerative processes elicited by SVD-related damage to the WM tracts that disrupt the connections between remote brain regions.^1, 24^ Indeed, in this study we also noted widespread alterations of WM tracts in relation to SVD burden, reflected by decreased FA, and increased MD, AD, and RD, all of which are established markers of WM microstructural damage in ageing and dementia.^19^ It is noteworthy that these associations remained after adding WMH masks as voxel-wise confounds, indicating that WMH alone did not explain these associations, but that SVD severity beyond WMH instead concurs with white matter microstructural abnormalities.^37^ These findings thus underscore that total SVD burden scores more comprehensively characterize the consequences of SVD than the sole consideration of single MRI features.^3, 4^ We also revealed that, relative to cognitively healthy adults, those with cognitive impairments (MoCA<26) had more pronounced negative associations between SVD and WM diffusivity, highlighting the central role of WM microstructure in the emergence of cognitive impairments.^31, 37^ Altogether, we demonstrate that the accumulation of SVD-related damage may manifest clinically through diffuse effects on WM tracts throughout the brain, and relate to gross changes in brain regions distant from the initial lesion site.^1, 24^

The importance of managing midlife vascular health to maintain cognition in late life has been demonstrated previously in the Whitehall II and other observational cohorts.^5-7, 9, 38^ Our findings suggest that SVD may also play an important part in this relationship. However, the mechanisms linking SVD-associated injuries with our observed changes in mid-life blood pressure, diffuse brain microstructural alterations, and subsequent cognitive dysfunction remain to be elucidated.^26^ One potential mediator in this pathway which could provide a direct link between peripheral and central vascular damage may be large artery stiffening, which has been associated with SVD burden^39^, midlife hypertension, damage to the blood-brain barrier, subsequent brain atrophy, and cognitive impairment.^38, 40^ Future longitudinal studies which include multiple assessments of SVD features would help unravel these complex mechanisms and temporal dynamics in further detail.

Strengths of the current study include the detailed examination of participants over a 25-year period, including repeated assessments of vascular risk and cognitive functions and the acquisition of advanced and multi-modal MRI scans in old age, allowing for a comprehensive characterization of total SVD burden. However, several limitations should be noted. First, the generalizability of our findings is limited as participants of the Whitehall II study are predominantly Caucasian, generally more educated, and only 20.7% individuals were female in this study. Second, in contrast to most of the previous work that investigated global SVD burden among patient groups (e.g., stroke or hypertensive populations),^3, 11, 28, 29^ our sample was relatively healthy, partially due to our selection criteria (e.g., MRI compatibility; no gross brain abnormalities). Moreover, as in other studies^3, 4, 10, 11^, the distribution of SVD burden in this sample was skewed, with low frequency of severe SVD. Together, this may have resulted in an underestimation of the strength of our observed associations and may in part explain why our observed effects were only small-to-medium. We therefore emphasize the need to replicate our findings in diverse cohort studies.

We demonstrated longitudinal associations of late-life SVD burden with vascular and cognitive functioning. Importantly, we observed that midlife blood pressure (mean age of 48) may contribute to SVD burden 20 years later (mean age 70). Furthermore, SVD burden related to slightly faster rates of cognitive decline, together with more pronounced and widespread differences in GM and WM microstructure. Together, our findings further emphasize the importance of midlife vascular health to maintain brain structure and function.

## Data Availability

The study follows Medical Research Council data sharing policies (https://mrc.ukri.org/research/policies-and-guidance-for-researchers/data-sharing/), and data from the Whitehall II Study and Whitehall II Imaging Sub-study are accessible via application to the Dementias Platform UK (https://portal.dementiasplatform.uk/).

## Acknowledgments

We thank all the participating civil service departments; the British Occupational Health and Safety Agency; the British Council of Civil Service Unions; all participating civil servants in the Whitehall II Study; and all members of the Whitehall II Study team at University College London who so helpfully collaborated with us. The Whitehall II Study team comprises research scientists, statisticians, study coordinators, nurses, data managers, administrative assistants, and data entry staff, who make the study possible. Staff at the Wellcome Centre for Integrative Neuroimaging in Oxford, in particular research radiographers Michael Sanders, MSc, Jon Campbell, MMRTech, BcAppSc, Caroline Young, DCR(R), David Parker, BSc(Hons), who acquired the scans. Martin R. Turner, MA, MBBS, PhD, FRCP (Wellcome Centre for Integrative Neuroimaging, Oxford, United Kingdom), and his colleagues advised on incidental findings and taking over clinical responsibility for such participants. In addition, we thank Charlotte L. Allan, Anya G. Topiwala, and Vyara Valkanova for the visual ratings of white matter hyperintensities. No compensation was provided for staff contributions to this study.

## Author’s contributions

**MGJ** and **SS** conceived the present study and analyzed the data. **MGJ** drafted the manuscript, created the figures, and supplementary materials. **MGJ, CEM, NF, EZs, ASM, MK, KPE** and **SS** contributed to the study concept and design. **NF** and **EZs** had a major role in acquisition of the data. **MGJ, LG, CEM, MA, LM, AMGdL, KW, FEdL, ASM, MK, KPE** and **SS** contributed to analysis and/or interpretation of the data. **CEM, ASM, MK, KPE** obtained study funding. All authors critically revised the manuscript for intellectual content and approved the final version of the manuscript.

## Supplemental material

Supplemental material for this article is available online.

## References

1. Ter Telgte A, van Leijsen EMC, Wiegertjes K, et al. Cerebral small vessel disease: from a focal to a global perspective. Nat Rev Neurol 2018; 14: 387-398. 2018/05/29. DOI:10.1038/s41582-018-0014-y.

2. Wardlaw JM, Smith EE, Biessels GJ, et al. Neuroimaging standards for research into small vessel disease and its contribution to ageing and neurodegeneration. The Lancet Neurology 2013; 12: 822–838. DOI:10.1016/s1474-4422(13)70124-8.

3. Staals J, Makin SD, Doubal FN, et al. Stroke subtype, vascular risk factors, and total MRI brain small-vessel disease burden. Neurology 2014; 83: 1228–1234. DOI:10.1212/WNL.0000000000000837.

4. Klarenbeek P, van Oostenbrugge RJ, Rouhl RP, et al. Ambulatory blood pressure in patients with lacunar stroke: association with total MRI burden of cerebral small vessel disease. Stroke 2013; 44: 2995–2999.

5. Suri S, Topiwala A, Chappell MA, et al. Association of midlife cardiovascular risk profiles with cerebral perfusion at older ages. JAMA network open 2019; 2: e195776–e195776. DOI:10.1001/jamanetworkopen.2019.5776.

6. Zsoldos E, Mahmood A, Filippini N, et al. Association of midlife stroke risk with structural brain integrity and memory performance at older ages: a longitudinal cohort study. Brain Communications 2020; 2. DOI:10.1093/braincomms/fcaa026.

7. Lane CA, Barnes J, Nicholas JM, et al. Associations between blood pressure across adulthood and late-life brain structure and pathology in the neuroscience substudy of the 1946 British birth cohort (Insight 46): an epidemiological study. The Lancet Neurology 2019; 18: 942–952. DOI:10.1016/S1474-4422(19)30228-5.

8. Livingston G, Huntley J, Sommerlad A, et al. Dementia prevention, intervention, and care: 2020 report of the Lancet Commission. The Lancet 2020; 396: 413–446. DOI:10.1016/S0140-6736(20)30367-6.

9. Muller M, Sigurdsson S, Kjartansson O, et al. Joint effect of mid-and late-life blood pressure on the brain: the AGES-Reykjavik study. Neurology 2014; 82: 2187–2195. DOI:10.1212/WNL.0000000000000517.

10. Field TS, Doubal FN, Johnson W, et al. Early life characteristics and late life burden of cerebral small vessel disease in the Lothian Birth Cohort 1936. Aging (Albany NY) 2016; 8:2039. DOI:10.18632/aging.101043.

11. Lau KK, Li L, Simoni M, et al. Long-Term Premorbid Blood Pressure and Cerebral Small Vessel Disease Burden on Imaging in Transient Ischemic Attack and Ischemic Stroke: Population-Based Study. Stroke 2018; 49: 2053–2060. DOI:10.1161/STROKEAHA.118.021578.

12. Blair GW, Hernandez MV, Thrippleton MJ, et al. Advanced neuroimaging of cerebral small vessel disease. Current treatment options in cardiovascular medicine 2017; 19:56. DOI:10.1007/s11936-017-0555-1.

13. Filippini N, Zsoldos E, Haapakoski R, et al. Study protocol: the Whitehall II imaging sub-study. BMC psychiatry 2014; 14:159. DOI:10.1186/1471-244X-14-159.

14. Marmot M and Brunner E. Cohort profile: the Whitehall II study. International journal of epidemiology 2005; 34: 251–256. DOI:10.1093/ije/dyh372.

15. D’Agostino RB, Wolf PA, Belanger AJ, et al. Stroke risk profile: adjustment for antihypertensive medication. The Framingham Study. Stroke 1994; 25: 40–43. DOI:https://doi.org/10.1161/01.STR.25.1.40.

16. Stewart AL, Mills KM, King AC, et al. CHAMPS physical activity questionnaire for older adults: outcomes for interventions. Med Sci Sports Exerc 2001; 33: 1126-1141. 2001/07/11. DOI:10.1097/00005768-200107000-00010.

17. Heim A. The AH4 group test of intelligence. Windsor: NFER-Nelson 1970.

18. Nasreddine ZS, Phillips NA, Bédirian V, et al. The Montreal Cognitive Assessment, MoCA: a brief screening tool for mild cognitive impairment. J Am Geriatr Soc 2005; 53: 695-699. 2005/04/09. DOI:10.1111/j.1532-5415.2005.53221.x.

19. Suri S, Topiwala A, Mackay CE, et al. Using structural and diffusion magnetic resonance imaging to differentiate the dementias. Current neurology and neuroscience reports 2014; 14:475. DOI:10.1007/s11910-014-0475-3.

20. Griffanti L, Jenkinson M, Suri S, et al. Classification and characterization of periventricular and deep white matter hyperintensities on MRI: A study in older adults. NeuroImage 2018; 170: 174–181. DOI:https://doi.org/10.1016/j.neuroimage.2017.03.024.

21. Singh-Manoux A, Kivimaki M, Glymour MM, et al. Timing of onset of cognitive decline: results from Whitehall II prospective cohort study. Bmj 2012; 344:d7622. DOI:https://doi.org/10.1136/bmj.d7622.

22. Elbaz A, Shipley MJ, Nabi H, et al. Trajectories of the Framingham general cardiovascular risk profile in midlife and poor motor function later in life: the Whitehall II study. International journal of cardiology 2014; 172: 96–102. DOI:10.1016/j.ijcard.2013.12.051.

23. Beck D, de Lange A-MG, Maximov II, et al. White matter microstructure across the adult lifespan: A mixed longitudinal and cross-sectional study using advanced diffusion models and brain-age prediction. NeuroImage 2021; 224:117441. DOI:https://doi.org/10.1016/j.neuroimage.2020.117441.

24. Wardlaw JM, Smith C and Dichgans M. Small vessel disease: mechanisms and clinical implications. The Lancet Neurology 2019; 18: 684–696. DOI:https://doi.org/10.1016/S1474-4422(19)30079-1.

25. Cannistraro RJ, Badi M, Eidelman BH, et al. CNS small vessel disease: a clinical review. Neurology 2019; 92: 1146–1156. DOI:10.1212/WNL.0000000000007654.

26. Østergaard L, Engedal TS, Moreton F, et al. Cerebral small vessel disease: capillary pathways to stroke and cognitive decline. Journal of Cerebral Blood Flow & Metabolism 2016; 36: 302–325. DOI:10.1177/0271678X15606723.

27. Debette S, Schilling S, Duperron M-G, et al. Clinical significance of magnetic resonance imaging markers of vascular brain injury: a systematic review and meta-analysis. JAMA neurology 2019; 76: 81–94. DOI:10.1001/jamaneurol.2018.3122.

28. Uiterwijk R, van Oostenbrugge RJ, Huijts M, et al. Total cerebral small vessel disease MRI score is associated with cognitive decline in executive function in patients with hypertension. Frontiers in aging neuroscience 2016; 8:301. DOI:10.3389/fnagi.2016.00301.

29. Al Olama AA, Wason JM, Tuladhar AM, et al. Simple MRI score aids prediction of dementia in cerebral small vessel disease. Neurology 2020; 94: e1294–e1302. DOI:https://doi.org/10.1212/wnl.0000000000009141.

30. Xu X, Hilal S, Collinson SL, et al. Association of magnetic resonance imaging markers of cerebrovascular disease burden and cognition. Stroke 2015; 46: 2808–2814. DOI:10.1161/STROKEAHA.115.010700.

31. Dichgans M and Leys D. Vascular cognitive impairment. Circulation research 2017; 120: 573–591. DOI:10.1161/CIRCRESAHA.116.308426.

32. Stern Y, Arenaza-Urquijo EM, Bartrés-Faz D, et al. Whitepaper: Defining and investigating cognitive reserve, brain reserve, and brain maintenance. Alzheimer’s & Dementia 2018. DOI:10.1016/j.jalz.2018.07.219.

33. Lambert C, Narean JS, Benjamin P, et al. Characterising the grey matter correlates of leukoaraiosis in cerebral small vessel disease. NeuroImage: Clinical 2015; 9: 194–205. DOI:10.1016/j.nicl.2015.07.002.

34. Lambert C, Benjamin P, Zeestraten E, et al. Longitudinal patterns of leukoaraiosis and brain atrophy in symptomatic small vessel disease. Brain 2016; 139: 1136–1151. DOI:10.1093/brain/aww009.

35. Jack Jr CR, Bennett DA, Blennow K, et al. NIA-AA research framework: toward a biological definition of Alzheimer’s disease. Alzheimer’s & Dementia 2018; 14: 535–562. DOI:10.1016/j.jalz.2018.02.018.

36. Sweeney MD, Montagne A, Sagare AP, et al. Vascular dysfunction—The disregarded partner of Alzheimer’s disease. Alzheimer’s & Dementia 2019; 15: 158–167. DOI:10.1016/j.jalz.2018.07.222.

37. Croall ID, Lohner V, Moynihan B, et al. Using DTI to assess white matter microstructure in cerebral small vessel disease (SVD) in multicentre studies. Clinical science 2017; 131: 1361–1373. DOI:10.1042/CS20170146.

38. Suri S, Chiesa ST, Zsoldos E, et al. Associations between arterial stiffening and brain structure, perfusion, and cognition in the Whitehall II Imaging Sub-study: A retrospective cohort study. PLoS medicine 2020; 17:e1003467. DOI:10.1371/journal.pmed.1003467.

39. Song T-J, Kim YD, Yoo J, et al. Association between aortic atheroma and cerebral small vessel disease in patients with ischemic stroke. Journal of stroke 2016; 18:312. DOI:10.5853/jos.2016.00171.

40. Pase MP, Himali JJ, Mitchell GF, et al. Association of aortic stiffness with cognition and brain aging in young and middle-aged adults: the Framingham Third Generation Cohort Study. Hypertension 2016; 67: 513–519. DOI:10.1161/HYPERTENSIONAHA.115.06610.

