## Supplemental Material for "Association of cerebral small vessel disease burden with brain structure and cognitive and vascular risk trajectories in mid-to-late life"

**Overview of supplementary materials**

*Supplementary methods*

*Description of cohort profile*

*Description of SVD ratings on MRI*

*MRI pre-processing steps*

*Study variables*

*Description of linear mixed effect models*

*Supplementary tables*

*Table S1. MRI acquisition parameters*

*Table S2. Participant characteristics of the Whitehall II study*

*Table S3. Included vs. complete sample of Whitehall II Imaging Sub-study*

*Supplementary figures*

*Figure S1. Flowchart of subject inclusion*

*Figure S2. Venn diagram of cerebral small vessel disease MRI score*

**Supplementary methods**

*Description of cohort profile*

The Whitehall II study targeted all civil servants that worked in the London offices of 20 Whitehall departments between 1985–1988, established by University College London. The initial Wave included 10 308 British Civil servants (6895 men), aged 35–55 years. Whitehall II Study participants have received detailed clinical follow-ups for up to 30 years at 5-year intervals (1991-1994, Wave 3; 1997-1999, Wave 5; 2002-2004, Wave 7; 2007-2009, Wave 9; 2012-2013, Wave 11); 2015-2016, Wave 12). Since the inception of the Whitehall II study, the retention rate for this cohort has been relatively high; about 87% of Wave 9 participants returned for the follow-up at Wave 11. The Whitehall II Imaging-Sub study randomly selected 774 participants aged 60-85 years from the Whitehall II Wave 11 cohort for multi-modal brain MRI work-up and cognitive tests at the University of Oxford. For the Imaging Sub-study, participants were included with contraindications to MRI scanning (e.g., particular metallic implants) or who were unable to travel to Oxford without assistance.[1, 2]

*Description of SVD ratings on MRI*

Periventricular and deep white matter hyperintensities (WMH) were rated by trained raters (C.L.A., A.G.T., V.V.; see acknowledgments) on FLAIR images using the Fazekas scale, providing a score between 0-3 depending on the severity of WMH in the corresponding brain areas.[3] Enlarged perivascular spaces (EPVS) and lacunes were rated by an experienced rater (M.G.J.) following extensive training, and in consensus with other experienced raters (S.S., L.M.). Lacunes were rated using both T1-weighted and FLAIR images, following established criteria to distinguish lacunes from EPVS.[4, 5] The intra-rater reliability for lacune ratings indicated high similarity, as reflected by an intraclass correlation (ICC) of 0.91, based on a random sample of 25 participants. EPVS were assessed in the basal ganglia on T1-weighted images using the validated qualitative EPVS rating scale, as the spatial resolution of T2-weighted images was not adequate to reliably detect EPVS.[6] The ICC for EPVS was 0.85, based on a random sample of 30 participants, indicating good intra-rater reliability. We used a semi-automatic detection method to identify possible cerebral microbleeds (CMBs), based on the radial symmetry transform.[7] Subsequently, one experienced rater (S.S.) evaluated all possible CMBs using previously established criteria[7, 8], and in consensus with a clinical psychiatrist (K.P.E.). The intra-rater reliability yielded excellent results (ICC = 0.92, based on a random sample of 100 participants).

*MRI pre-processing steps*

MRI scans were analysed using FMRIB Software Library v6.0 (FSL; https://fsl.fmrib.ox.ac.uk/).[9]

**T1 images** were bias corrected, brain extracted using FSL-ANAT and segmented using FSL-FAST to provide estimates of grey matter (GM, white matter (WM) and total brain volume (TBV).[10]

**Diffusion-weighted images** were pre-processed using FMRIB's diffusion toolbox.[11] Briefly, after applying motion and eddy current corrections with FSL-TOPUP, diffusivity maps for each metric was extracted using DTIFit and aligned into standard space using FMRIB's Nonlinear Registration Tool (FNIRT).

**FLAIR scans** were used to extract WMH using the Brain Intensity AbNormality Classification Algorithm (BIANCA).[12] This algorithm uses both intensity features (provided from FLAIR, T1 images, and fractional anisotropy) and spatial features to classify all voxels. BIANCA was initially trained on manually segmented WMH masks of participants who were scanned on the Prisma scanner (N = 24), Verio scanner (N=24), and an independent sample from the UK Biobank Study (N = 12) to avoid scanner-dependent bias effects.

Additional information on pre-processing was mentioned previously.[1, 13]

*Study variables*

The **FSRS** was based on age, sex, systolic blood pressure, use of antihypertensive medications, diabetes mellitus, current smoking, current or history of atrial fibrillation, left ventricular hypertrophy, and current or history of cardiovascular disease.[14] These measurements were obtained using both questionnaires and clinical assessments, using standard operating protocols, as described previously.[15, 16] **Blood pressure** measurements were obtained in sitting position after five minutes rest; the average of two measurements was used for further analysis. The use of **antihypertensive medication** was self-reported (e.g., diuretics, beta blockers, angiotensin-converting enzyme inhibitors, and calcium channel blockers). **Diabetes** was defined by having a fasting glucose level of ≥ 7.0 mml/L or a 2hr post-load glucose level of ≥ 11.1 mml/L, based on glucose measurements obtained from venous blood; self-reported diabetes diagnosed by a doctor or use of diabetes medication.[17] **Smoking behaviour** was self-reported (current, past/no smoking). A standard electrocardiogram analysis combined with manual review and Minnesota code classification system for electrocardiographic findings was used to identify **atrial fibrillation** and **left ventricular hypertrophy**.[18] **Cardiovascular disease** was evaluated using corroborated records from the general practitioner, hospital, and electrocardiogram and angiogram examinations at Wave 1, 3 and 5. Subsequently, the FSRS was computed using the beta coefficients of the Cox proportional hazards regression model in the Framingham Study, to indicate an individual's 10-year risk of stroke.[19]

The longitudinal test battery of the Whitehall II cohort includes several cognitive tests, proven to be sensitive to detect changes in cognitive functions in this study population.[20] The complete test battery took 30 minutes to complete. To measure **letter fluency**, participants were instructed to recall as many words beginning with an "S" within one minute. For **semantic fluency**, participants were given similar instructions, but instead needed to recall as many animal names. **Short-term memory** was evaluated by initially presenting a list of 20 one or two syllable words at two seconds intervals. Subsequently, participants were asked to recall as many of the word list, within two minutes. The Alice Heim 4-I test composes 65 verbal and mathematical reasoning items with increasing difficulty (e.g., where participants had to identify certain patterns or rules), covering **verbal and**

**numerical reasoning**.[21] Participants were given 10 minutes to complete the test. Besides this, the test battery also included the Mill Hill vocabulary test[22] and the Mini Mental State Examination[23], however these tests were not included in the present study due to the observed ceiling effects.

Additional information on the vascular and cognitive study variables was mentioned previously.[20, 24-26]

*Description of linear mixed effect models*

To investigate the association between the cerebral small vessel disease (SVD) MRI score and trajectories of vascular risk and cognitive performance over 25 years, we employed the following equation for each dependent variable of interest (V):

$$V_{ij} = \beta_0 + \beta_1 time_{ij} + \beta_2 time_{ij}^2 + \beta_3 SVD_{ij} + \beta_4 SVD_{ij} time_{ij} + \beta_5 SVD_{ij} time_{ij}^2 + \beta_6 X_{1i} + \beta_7 X_{2i} + \beta_8 X_{2i} time_{ij} + \beta_9 X_{1i} time_{ij}^2 + U_{0i} + U_{1i} time_{ij} + e_{ij}$$

$V_{ij}$ is the dependent variable of interest of the $i^{th}$ participant at the $j^{th}$ occasion,
$time_{ij}$ is years since the baseline measurement between 1995-1999 for the $i^{th}$ participant at the $j^{th}$ occasion,
$time^2_{ij}$ is the orthogonalized polynomial quadratic time term for the $i^{th}$ participant at the $j^{th}$ occasion,
$SVD_i$ is the total SVD score obtained during the Whitehall Imaging Sub-study of the $i^{th}$ participant,
$X_{1i}$ is the covariate for scanner model (Prisma vs. Verio) for the $i^{th}$ participant,
$X_{2i}$ is a vector of covariates (age at baseline, sex, education) for the $i^{th}$ participant,
$U_{0i}$ is the random intercept, $U_{1i}$ is the random slope, and $e_{ij}$ is the residual.

The dependent variables of interest focused on vascular risk and cognitive performance.

Vascular risk was defined using the Framingham Stroke Risk Score (FSRS) and mean arterial pressure (MAP). Due to the skewed distribution of the residuals, FSRS was log-transformed.

Cognitive performance was covered for the following domains: letter fluency, semantic fluency, verbal reasoning, numerical reasoning, and global cognition.

Vascular risk factors were measured at six waves (j= 1, 2, 3, 4, 5, 6), whereas measures for cognitive performance were included from five waves (j = 1, 2, 3, 4, 5).

Parameter estimates were obtained with the Maximum Likelihood method from the *nlme* in R version 3.6.1.

Main effects of SVD burden are indicative of whether SVD burden scores (1-3) differed from the reference score (no burden; 0) on the variable of interest at baseline. The interaction of SVD burden with time indicates whether a higher SVD burden (1-3) is associated with different longitudinal trajectories (i.e., slopes) of the respective variable of interest as compared to the reference score. To allow for individual rates of change of the dependent variables over time for each participant, we fitted the intercept and slope as random effects.

We implemented a continuous autoregressive moving-average correlation structure to consider repeated measures for each individual.

**Supplementary tables**

**Table S1.** MRI acquisition parameters

| | | TR (ms) | TE (ms) | TI (ms) | Flip angle (°) | Field of view (mm) | Matrix (voxels) |
|---|---|---|---|---|---|---|---|
| **T1-weighted** | | | | | | | |
| | Verio | 2530 | 1.79/3.65/ 5.51/7.37 | 1380 | 7 | 256 | 1.0x1.0x1.0 |
| | Prisma | 1900 | 3.97 | 904 | 8 | 192 | 1.0x1.0x1.0 |
| **FLAIR** | | | | | | | |
| | Verio | 9000 | 73 | 2500 | 150 | 220 | 0.9x0.9x3.0 |
| | Prisma | 9000 | 73 | 2500 | 150 | 220 | 0.4x0.4x3.0 |
| **T2*-weighted** | | | | | | | |
| | Verio | 36 | 30 | - | 15 | 220 | 0.7x0.7x1.5 |
| | Prisma | 1230 | 13.4 | - | 25 | 206 | 0.8x0.8x5.0 |
| **DWI** | | | | | | | |
| | Verio | 8900 | 91.2 | - | - | 192 | 2.0x2.0x2.0 |
| | Prisma | 8900 | 91 | - | - | 192 | 2.0x2.0x2.0 |
| **B0** | | | | | | | |
| | Verio | 8900 | 91.2 | - | - | 192 | 2.0x2.0x2.0 |
| | Prisma | 8900 | 91 | - | - | 192 | 2.0x2.0x2.0 |

Abbreviations: FLAIR: fluid-attenuated inversion recovery; DWI: diffusion-weighted imaging; TR: repetition time; TE: echo time; TI: inversion time.

**Table S2.** Sample characteristics for the 623 participants of the Whitehall II study

| | **Wave 3** (1991-1994) | | **Wave 5** (1997-1999) | | **Wave 7** (2002-2004) | | **Wave 9** (2007-2009) | | **Wave 11** (2012-2013) | | **Wave 12** (2015-2016) | |
|---|---|---|---|---|---|---|---|---|---|---|---|---|
| **General characteristics** | N | Mean (SD or median (IQR) | N | Mean (SD or median (IQR) | N | Mean (SD or median (IQR) | N | Mean (SD or median (IQR) | N | Mean (SD or median (IQR) | N | Mean (SD or median (IQR) |
| Age | 623 | 47.81 (5.23) | 608 | 53.58 (5.22) | 609 | 59.07 (5.20) | 620 | 64.01 (5.21) | 623 | 68.10 (5.22) | 610 | 71.30 (5.23) |
| **Vascular risk factors** | | | | | | | | | | | | |
| MAP | 608 | 91.63 (9.71) | 585 | 90.64 (11.24) | 603 | 90.08 (10.90) | 619 | 87.94 (10.66) | 623 | 88.64 (10.46) | 606 | 89.19 (10.78) |
| BMI | 607 | 24.80 (3.32) | 517 | 25.26 (3.70) | 603 | 26.21 (3.91) | 620 | 26.29 (4.10) | 623 | 26.28 (4.11) | 606 | 26.39 (4.09) |
| FSRS | 588 | 3 (3-4) | 869 | 3 (3-4) | 594 | 4 (3-6) | 603 | 5 (4-7) | 602 | 6 (5-10) | 578 | 7 (5-11) |
| **Cognitive performance** | | | | | | | | | | | | |
| Letter fluency | 290 | 17.84 (4.17) | 560 | 17.72 (4.19) | 593 | 16.87 (3.81) | 615 | 16.13 (3.64) | 622 | 16.09 (4.01) | 602 | 15.58 (4.28) |
| Semantic fluency | 290 | 16.69 (3.44) | 559 | 17.21 (4.00) | 596 | 16.56 (3.59) | 617 | 15.93 (3.58) | 622 | 15.77 (3.59) | 602 | 15.80 (3.54) |
| Verbal reasoning | 289 | 25.17 (4.49) | 561 | 25.30 (4.23) | 596 | 24.10 (4.35) | 618 | 23.79 (4.58) | 622 | 23.72 (4.59) | 603 | 23.98 (4.77) |
| Numerical reasoning | 289 | 24.44 (5.28) | 561 | 24.39 (5.16) | 596 | 23.31 (5.11) | 618 | 22.89 (5.42) | 622 | 22.75 (5.53) | 603 | 21.84 (5.64) |
| Memory | 288 | 6.15 (2.15) | 558 | 7.31 (2.30) | 595 | 7.35 (2.17) | 618 | 6.59 (2.19) | 622 | 6.58 (2.24) | 599 | 5.68 (2.20) |

Abbreviations: SD: standard deviation; IQR: interquartile range; MAP: mean arterial pressure; FSRS: Framingham Stroke Risk Score.

**Table S3.** Included vs. complete sample of Whitehall II Imaging Sub-study at MRI wave

| | Included sample | Complete sample | P-value |
|---|---|---|---|
| Number of participants | 623 | 775 | |
| Age, mean (SD) | 69.96 (5.18) | 69.81 (5.19) | 0.59 |
| Female, N (%) | 129 (21%) | 150 (19%) | 0.58 |
| Education, mean (SD) | 14.11 (3.05) | 14.06 (3.06) | 0.74 |
| MoCA, median (IQR) | 28 (26-29) | 28 (26-29) | 0.74 |
| MAP, mean (SD) | 98.84 (11.82) | 98.79 (11.70) | 0.95 |
| BMI, mean (SD) | 25.96 (4.14) | 26.15 (4.17) | 0.38 |

Abbreviations: MoCA: Montreal cognitive assessment. MAP: mean arterial pressure. BMI: body mass index. Differences in characteristics were compared using independent *t*-test, Chi-square test or Mann-Whitney U test where appropriate. Data on MAP was missing in 4 (0.05%) from the complete sample.

**Supplementary figures**

**Figure S1.** Flowchart of subject inclusion

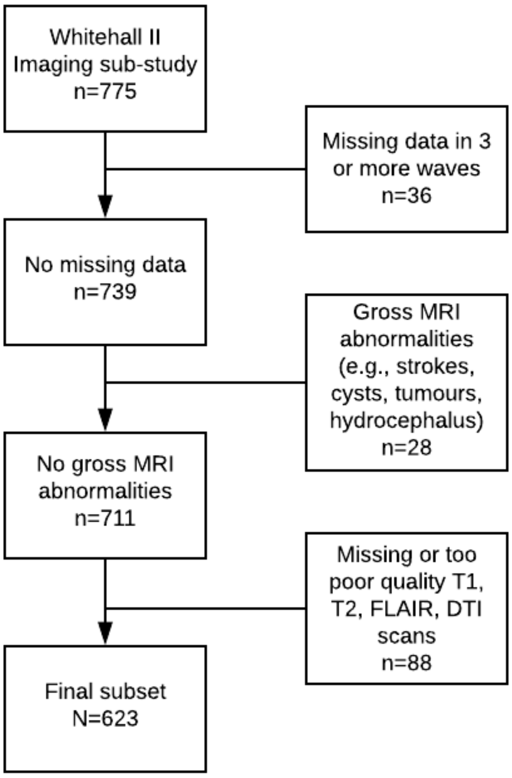

**Figure S2.** Venn diagram of cerebral small vessel disease MRI score

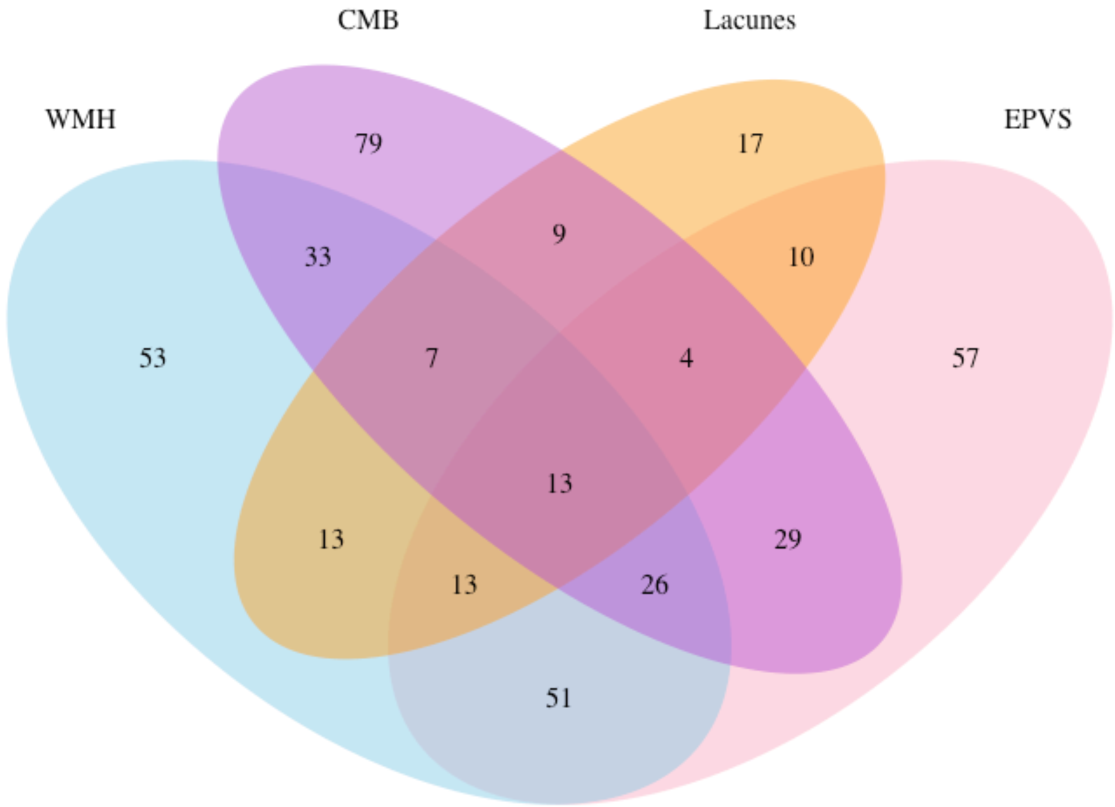


Abbreviations: WMH: white matter hyperintensities; CMB: cerebral microbleeds; EPVS: enlarged perivascular spaces. Numbers depict how many participants were within a certain category. Only 13 participants demonstrated all four features of cerebral small vessel disease on MRI.

**Supplemental references**

1. Filippini N, Zsoldos E, Haapakoski R, et al. Study protocol: the Whitehall II imaging sub-study. *BMC psychiatry* 2014; 14: 159. DOI: 10.1186/1471-244X-14-159.

2. Marmot M and Brunner E. Cohort profile: the Whitehall II study. *International journal of epidemiology* 2005; 34: 251-256. DOI: 10.1093/ije/dyh372.

3. Fazekas F, Chawluk JB, Alavi A, et al. MR signal abnormalities at 1.5 T in Alzheimer's dementia and normal aging. *American Journal of Roentgenology* 1987; 149: 351-356. DOI: 10.2214/ajr.149.2.351.

4. Duering M, Csanadi E, Gesierich B, et al. Incident lacunes preferentially localize to the edge of white matter hyperintensities: insights into the pathophysiology of cerebral small vessel disease. *Brain* 2013; 136: 2717-2726.

5. Benjamin P, Trippier S, Lawrence AJ, et al. Lacunar infarcts, but not perivascular spaces, are predictors of cognitive decline in cerebral small-vessel disease. *Stroke* 2018; 49: 586-593.

6. Potter GM, Chappell FM, Morris Z, et al. Cerebral perivascular spaces visible on magnetic resonance imaging: development of a qualitative rating scale and its observer reliability. *Cerebrovascular diseases* 2015; 39: 224-231.

7. Kuijf HJ, Brundel M, de Bresser J, et al. Semi-automated detection of cerebral microbleeds on 3.0 T MR images. *PLoS One* 2013; 8: e66610.

8. Linn J. Imaging of cerebral microbleeds. *Clinical neuroradiology* 2015; 25: 167-175.

9. Jenkinson M, Beckmann CF, Behrens TE, et al. Fsl. *Neuroimage* 2012; 62: 782-790.

10. Zhang Y, Brady M and Smith S. Segmentation of brain MR images through a hidden Markov random field model and the expectation-maximization algorithm. *IEEE transactions on medical imaging* 2001; 20: 45-57.

11. Smith SM, Jenkinson M, Johansen-Berg H, et al. Tract-based spatial statistics: voxelwise analysis of multi-subject diffusion data. *Neuroimage* 2006; 31: 1487-1505.

12. Griffanti L, Jenkinson M, Suri S, et al. Classification and characterization of periventricular and deep white matter hyperintensities on MRI: A study in older adults. *NeuroImage* 2018; 170: 174-181. DOI: https://doi.org/10.1016/j.neuroimage.2017.03.024.

13. Suri S, Chiesa ST, Zsoldos E, et al. Associations between arterial stiffening and brain structure, perfusion, and cognition in the Whitehall II Imaging Sub-study: A retrospective cohort study. *PLoS medicine* 2020; 17: e1003467. DOI: 10.1371/journal.pmed.1003467.

14. Wolf PA, D'Agostino RB, Belanger AJ, et al. Probability of stroke: a risk profile from the Framingham Study. *Stroke* 1991; 22: 312-318.

15. Kaffashian S, Dugravot A, Brunner EJ, et al. Midlife stroke risk and cognitive decline: a 10-year follow-up of the Whitehall II cohort study. *Alzheimer's & Dementia* 2013; 9: 572-579. DOI: https://doi.org/10.1016/j.jalz.2012.07.001.

16. Kivimäki M, Shipley MJ, Allan CL, et al. Vascular risk status as a predictor of later-life depressive symptoms: a cohort study. *Biological psychiatry* 2012; 72: 324-330.

17. Association AD. Report of the expert committee on the diagnosis and classification of diabetes mellitus. *Diabetes care* 2003; 26: s5-s20. DOI: https://doi.org/10.2337/diacare.26.2007.S5.

18. Blackburn H, Prineas R and Crow R. The Minnesota Code: manual for electrocardiographic findings: standards and procedures for measurement and classification. *Littleton, Mass, Wright* 1982.

19. D'Agostino RB, Wolf PA, Belanger AJ, et al. Stroke risk profile: adjustment for antihypertensive medication. The Framingham Study. *Stroke* 1994; 25: 40-43. DOI: https://doi.org/10.1161/01.STR.25.1.40.

20. Singh-Manoux A, Kivimaki M, Glymour MM, et al. Timing of onset of cognitive decline: results from Whitehall II prospective cohort study. *Bmj* 2012; 344: d7622. DOI: https://doi.org/10.1136/bmj.d7622.

21. Heim A. The AH4 group test of intelligence. *Windsor: NFER-Nelson* 1970.

22. Raven JC. *Guide to Using the Mill Hill Vocabulary Scale with Progressive Matrices (1938)*. HK Lewis, 1954.

23. Folstein MF, Folstein SE and McHugh PR. “Mini-mental state”: a practical method for grading the cognitive state of patients for the clinician. *Journal of psychiatric research* 1975; 12: 189-198.

24. Singh-Manoux A, Britton AR and Marmot M. Vascular disease and cognitive function: evidence from the Whitehall II Study. *Journal of the American Geriatrics Society* 2003; 51: 1445-1450.

25. Zsoldos E, Mahmood A, Filippini N, et al. Association of midlife stroke risk with structural brain integrity and memory performance at older ages: a longitudinal cohort study. *Brain Communications* 2020; 2. DOI: 10.1093/braincomms/fcaa026.

26. Suri S, Topiwala A, Chappell MA, et al. Association of midlife cardiovascular risk profiles with cerebral perfusion at older ages. *JAMA network open* 2019; 2: e195776-e195776. DOI: 10.1001/jamanetworkopen.2019.5776.
